# Work Status, Functional Recovery, and Quality of Life of Cardiac Arrest Survivors After Hospital Discharge

**DOI:** 10.1101/2023.08.29.23294804

**Authors:** Yi-Wen Wu, Tai-Yuan Chen, Chien-Hua Huang, Yu-Jen Chu, Wei-Ting Chen, Kuan-Ting Lu, Wei-Tien Chang, Hooi-Nee Ong, Wen-Jone Chen, Min-Shan Tsai

**Author notes:** **Name and Address of Corresponding Authors:** Min-Shan Tsai, MD, PhD Department of Emergency Medicine, National Taiwan University Hospital No. 7, Zhongshan S. Road, Taipei, Taiwan 100 |. The study was performed in National Taiwan University Hospital. Short running title: Recovery in cardiac arrest survivors.

## Abstract

**Background:** To investigate the work status, neurological recovery, and quality of life of cardiac arrest survivors within 1 year after hospital discharge.

**Methods:** A retrospective single center study included 71 non-traumatic adult cardiac arrest patients between 2017 and 2020, who survived more than 1 year after hospital discharge and agreed to participate the study. Questionnaire interviews through telephone visits were conducted with the enrolled patients, assessing their work status, neurological recovery, and quality of life at 3 months and 1 year after hospital discharge. Additionally, their use of medical services was also collected.

**Results:** Of the enrolled patients, 67 (94.4%) had a cerebral performance category (CPC) of 1 at discharge and the majority of patients (90.1%) returned home. Sixty-seven patients (94.4%) returned for outpatient visits at a median time of 6 days, 23 patients (32.4%) visited the ER at a median time of 74 days, and 22 patients (31.0%) were readmitted to the hospital at a median time of 58 days. In terms of mobility, self-care, usual activities, pain/discomfort, and health state assessed using the EQ-5D-5L, a significant decrease in impairment was observed from 3 months to 1 year. Moreover, patients demonstrated improved work status as well as improved scores for overall quality of life, general health, physical, psychological, social relations, and environmental status evaluated using the WHOQOL-BREF.

**Conclusions:** Within 1 year following hospital discharge, a considerable number of cardiac arrest survivors require medical support. However, a continual improvement in work status and quality of life were observed.

## Introduction

Advances in cardiopulmonary resuscitation (CPR) have led to a considerable increase in the successful rate of return of spontaneous circulation (ROSC) following emergency treatment.^1^, ^2^ The post-cardiac arrest syndrome including damage to the brain and heart after cardiac arrest contributes to a relatively high mortality rate in the early post-arrest period. With the introduction of post-arrest care and targeted temperature management (TTM),^3^, ^4^ the survival rate of patients with cardiac arrest continues to improve, and optimizing the quality of life and functional outcomes for these survivors after discharge becomes essential. In response to this requirement, the updated Advanced Cardiovascular Life Support guidelines emphasizes the importance of recovery in the chain of survival,^5^ which is a significant concern for both the families of survivors and the entire health-care system.

Hypoxia and subsequent ischemia-reperfusion injury, along with various sequelae, may influence physical, cognitive, and psychosocial aspects in cardiac arrest survivors.^6^ To evaluate the physical and mental well-being of these individuals after discharge, numerous studies have employed various self-assessment scales for quantifying quality of life and psychological stress.^3, 7-10^ Studies have demonstrated that cognitive impairment and emotional problems have a substantial impact on patients’ daily functioning, ability to return to work, and overall quality of life. Those with poor recovery often experience low participation, fatigue, depression, and mobility issues.^3^, ^8^ Additionally, patients with a diagnosis of depression or anxiety have a higher long-term mortality rate than those without such conditions.^10^ The prevalence of depression, anxiety, and other mental illnesses after out-of-hospital cardiac arrest (OHCA) can affect patients’ health-related quality of life.^9^

ROSC, survival, and neurological outcomes at hospital discharge are well-established and acceptable outcome indicators in cardiac arrest research.^11^ With advances in post-arrest care, an increasing number of survivors are being successfully discharged, and these clinical indicators alone may not fully capture the overall recovery and challenges faced by survivors after hospital discharge. Moreover, evaluations conducted at the time of discharge often do not adequately consider the challenges faced by survivors when they return home.^11^ There is a lack of data concerning the medical service requirement, work status and quality of life of cardiac arrest survivors after hospital discharge in Asian countries. Whether the work status and quality of life of cardiac arrest survivors would continuously improve also remains unknown. Therefore, the current study aimed to evaluate the use of medical service, work status, neurological recovery, and quality of life of cardiac arrest survivors within 1 year after hospital discharge.

## Methods

### Study Design

This retrospective study was conducted in National Taiwan University Hospital (NTUH), a tertiary referral medical centers with approximately 100,000 emergency department (ER) visits per year in Taipei city, Taiwan. The study protocol was approved by the Institutional Review Board of NTUH (202107138RIND).

### Study Population

Initially, 169 adults with non-traumatic cardiac arrest who survived to hospital discharge between 2017 and 2020 were recruited. The following patients were excluded: patients who died within 1 year following hospital discharge (n=42), those who could not be contacted after hospital discharge (n=35), and those who declined participation in the study (n=21). Ultimately, a total of 71 patients were included in this study (Figure 1).

**Figure 1.**
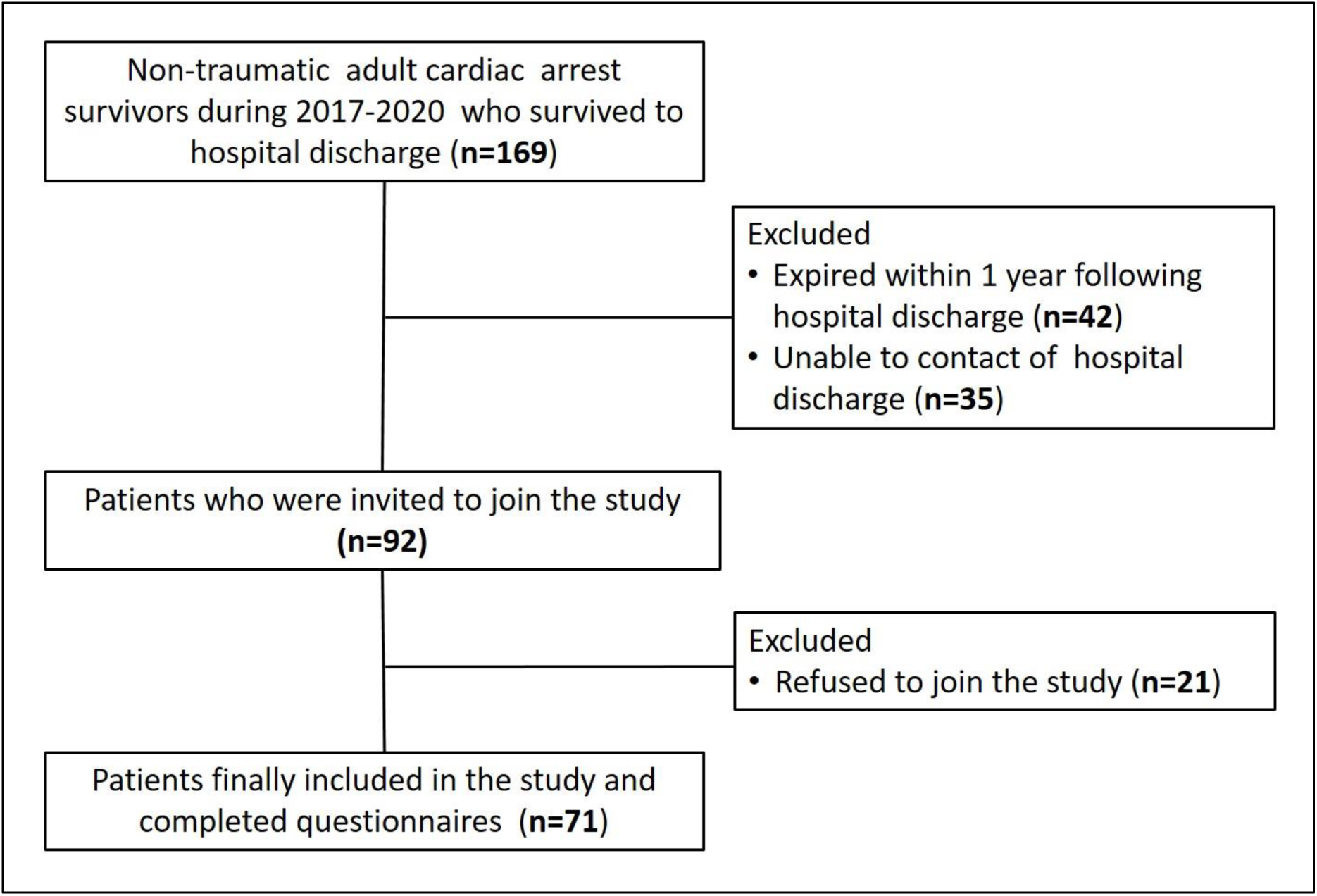
Flowchart of patient enrollment

Telephone visits and questionnaire interviews were conducted for the enrolled patients, which included assessments of their work status, neurological recovery, and quality of life at 3 months and 1 year after hospital discharge. Work status was defined as being employed in a salaried position and capable of performing tasks independently without requiring assistance from others. Neurological function was evaluated using Glasgow–Pittsburgh cerebral performance category (CPC) scores. CPC scores of 1–2 were defined as favorable neurologic outcomes and scores of 3–5 as poor neurologic outcomes. Quality of life was assessed using two questionnaires: the EuroQol 5-Dimension, 5-Level Health Scale (EQ-5D-5L) and World Health Organization Quality of Life-Brief (WHOQOL-BREF). The EQ-5D-5L evaluates individuals’ daily activities and provides insights into their health-related quality of life in terms of five dimensions.^12^, ^13^ The WHOQOL-BREF is a widely recognized instrument that surpasses the conventional assessment of physical and psychological aspects of health-related quality of life. It also incorporates social relationships and environment, making it a comprehensive tool for evaluating the overall quality of life.^14^ Both the EQ-5D-5L and WHOQOL-BREF were common tools for assessing affective and psychosocial well-being recommended by the American Heart Association.^11^

### Variables of Cardiac Arrest

This study collected baseline characteristics and data on resuscitation events, post-arrest care, and neurological outcomes of patients during their index admission from medical records. Initial shockable rhythm was defined as the initial cardiac rhythm recorded as ventricular fibrillation or ventricular tachycardia. A patient who experienced a recurrent cardiac arrest event within 1 h after ROSC was considered to have undergone repeated CPR. Cardiac arrest was considered to have a cardiac cause when ischemic heart disease, structural heart disease, acute coronary syndrome, heart failure, and arrhythmia without electrolyte imbalance were identified.^15^ TTM was applied to patients, with a target temperature of either 33 °C or 36 °C for 24 h. Extracorporeal membrane oxygenation (ECMO) applied within 7 days following cardiac arrest was recorded in this study. Emergent coronary angiography (CAG) was defined as CAG performed within 24 h after ROSC.^16^

This study recorded whether patients visited the outpatient department (OPD) or ER or were readmitted to the hospital after discharge. The diagnoses obtained during ER visits and hospital admissions were divided into several categories: cardiovascular (e.g., congestive heart disease, coronary artery disease, installation of an implantable cardioverter defibrillator, atrial fibrillation, ischemic cardiomyopathy, and chest pain), pulmonary (e.g., pneumonia and respiratory failure), infection (e.g., fever, cellulitis, and bacteremia), neurologic (e.g., thalamic hemorrhage), gastrointestinal (e.g., gastrointestinal symptoms and peptic ulcer diseases), endocrine/metabolism (e.g., hyperthyroidism and diabetic ketoacidosis), and miscellaneous (e.g., bruise, laceration wound, pancreatitis, and cholecystitis). Two physicians independently determined whether the diagnosis was related to the cause of the OHCA. In case of differing opinions between the two physicians, a third physician made the final judgment. Additionally, this study assessed the days alive and out of the hospital (DAOH), which was defined as the number of days during which the patient required no hospitalization and remained out of the hospital from the time of discharge up to 1 year.^17^ EuroQol 5-Dimension, 5-Level Health Scale (EQ-5D-5L)

The EQ-5D-5L, developed by the EuroQol Group, is widely used for assessing health-related quality of life (HRQoL) worldwide.^18^ The official EQ-5D-5L Taiwanese version in traditional Chinese characters was used in the current study (Supplementary Table 1).^13^ The questionnaire comprises five dimensions that capture an individual’s subjective perception of their health in the following areas: mobility, self-care, personal activities, pain/discomfort, and anxiety/expression. Responses are recorded on a 5-point scale, indicating levels of severity. In this study, patients were asked to rate their overall health status on a scale ranging from 0 (indicating the worst health imaginable) to 100 (indicating the best health imaginable).

In the current study, for each dimension in the EQ-5D-5L questionnaire, a score of ≥3 points was considered to indicate “more than moderate impaired,” and the health status scores of ≤60 were considered to indicate “more than moderate impaired.”

### World Health Organization Quality of Life-Brief (WHOQOL-BREF)

The World Health Organization Quality of Life-Brief-Taiwanese version [WHOQOL-BREF(TW)] has been adapted for Taiwanese culture and was used in the current study(Supplementary Table 2).^19^ This adaptation comprises two additional items (No.27 and No.28), resulting in a total of 28 items compared with the number of items in the original version. Of the questions for 28 items, two questions evaluating the overall condition are not included. The remaining 26 questions can be divided into four domains: Physical, comprising a total of 7 questions; psychological, comprising a total of 6 questions; social relationships, comprising a total of 4 questions; and environment, comprising a total of 9 questions. The item scores range from 1 (the worst condition) to 5 (the best condition), with the exception of three items (No.3, No.4, and No.26), which are reversely coded.

In the current study, all the reverse-coded questions in the WHOQOL-BREF(TW) were reversed again to obtain a higher score indicating a higher quality of life. To calculate the scores for each domain, the average scores of all the questions within each domain were multiplied by 4, resulting in domain scores ranging from 4 to 20.^20^

### Statistical Data Analyses

Categorical variables between groups were compared using the chi-squared test and are expressed as numbers and percentages. Continuous variables were compared between groups by using a Mann Whitney U test and are expressed as medians with quartiles. The repeat measurement was used to compare continuous variables between 3 months and 1 year. A p value of <0.05 was considered statistically significant. All statistical analyses were performed using SPSS Statistics for Windows, version 16.0 (SPSS Inc., Chicago, IL, USA).

## Results

Table 1 presents the baseline characteristics, CPR events, post-arrest care, and outcomes of the enrolled patients. Of the 71 patients included in the study, 59 (83.1%) were male, and the median age was 59 years. In this study sample, 42 (59.2%) had OHCA, 67 (94.4%) had a witnessed collapse, and 23 (32.4%) regained ROSC before arriving at the ER. Initial shockable cardiac rhythm was recorded in 44 patients (62.0%), and 58 patients (81.7%) were presumed to have a cardiac origin of cardiac arrest. Among the enrolled patients, 64 (90.1%) were discharged to their homes, 4 (5.6%) were transferred to another hospital for further care, and 3 (4.2%) were sent to a nursing home for specialized support after hospital discharge.

**Table 1.**
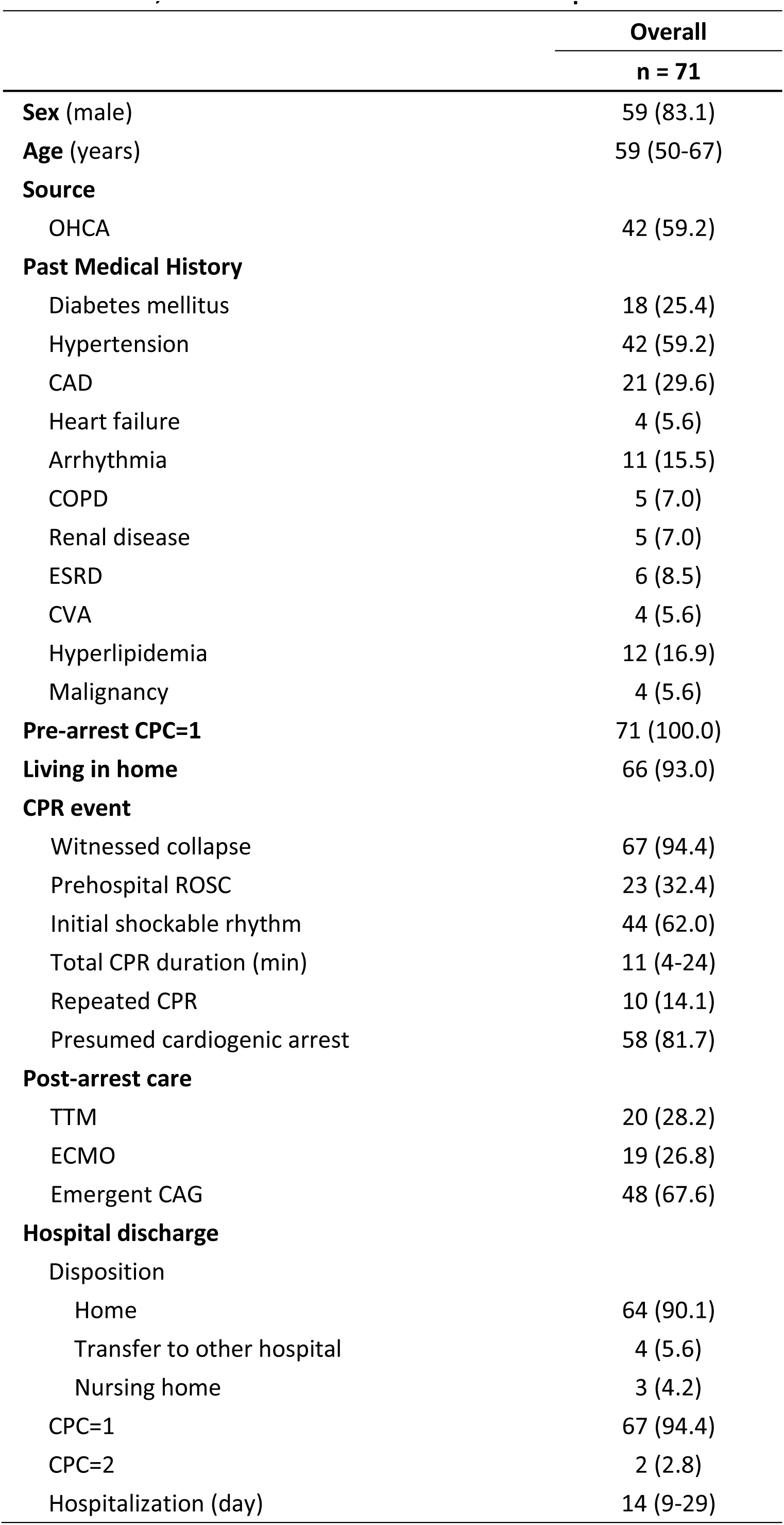
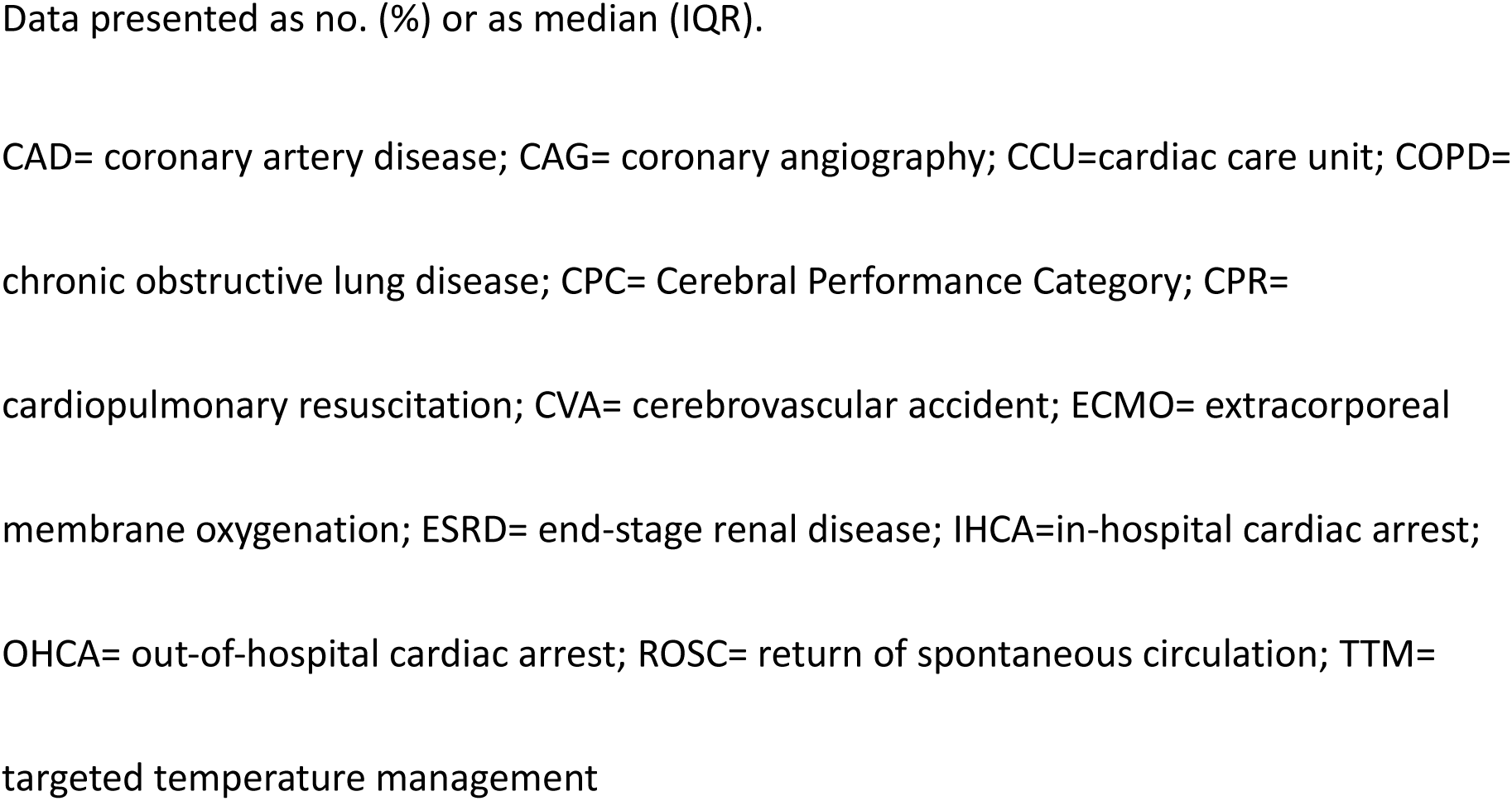
Characteristics, CPR events and outcomes of enrolled patients.

Table 2 presents the medical service requirements of the enrolled patients after hospital discharge. Most patients (n=67, 94.4%) returned to the OPD after discharge, with a median duration of 6 days from discharge to OPD visit. Twenty-three patients (32.4%) ever visited the ER after discharge, with a median duration of 74 days. Among them, 12 patients (16.9%) first visited the ER within 90 days after discharge. The diagnoses during ER visits were classified as follows: cardiovascular (n=8, 34.8%), pulmonary (n=2, 8.7%), infection (n=3, 13.0%), neurologic (n=1, 4.3%), gastrointestinal (n=2, 8.7%), endocrine/metabolism (n=2, 8.7%), and miscellaneous (n=5, 21.7%). In 9 patients (39.1%), the diagnoses during ER visits were considered related to the cause of cardiac arrest. Twenty-two patients (31.0%) were ever admitted to the hospital, with a median duration of 58 days after hospital discharge. Among them, 12 patients (16.9%) were admitted to the hospital within 90 days after discharge. Cardiovascular etiology was the main reason for hospital admission (n=13, 59.1%), and during hospitalization, most etiologies were considered to be related to the cause of cardiac arrest (n=14, 63.6%). Six patients (27.3%) were hospitalized after an ER visit. The median DAOH for the enrolled patients within 1 year following discharge was 361 days. A total of 32 patients (45.0%) ever visited the ER or were admitted during the first year following hospital discharge. The enrolled patients were further divided into cardiac (n=58) and non-cardiac (n=13) groups based on the cause of their cardiac arrest. The patients in the cardiac group visited the ER at a median duration of 62 days and were admitted to the hospital for a median duration of 37 days, which were earlier than the time points in the non-cardiac group. However, the analysis did not reveal a significant difference, which may be due to the limited number of patients. The diagnoses during ER visits and hospitalizations in the cardiac group also tended to be more correlated with the cause of cardiac arrest than the non-cardiac group (p=0.075). The cardiac group had more ER visits and included a higher number of patients visiting the ER compared with the non-cardiac group. Similar findings were observed for hospitalization (Figure 2).

**Figure 2.**
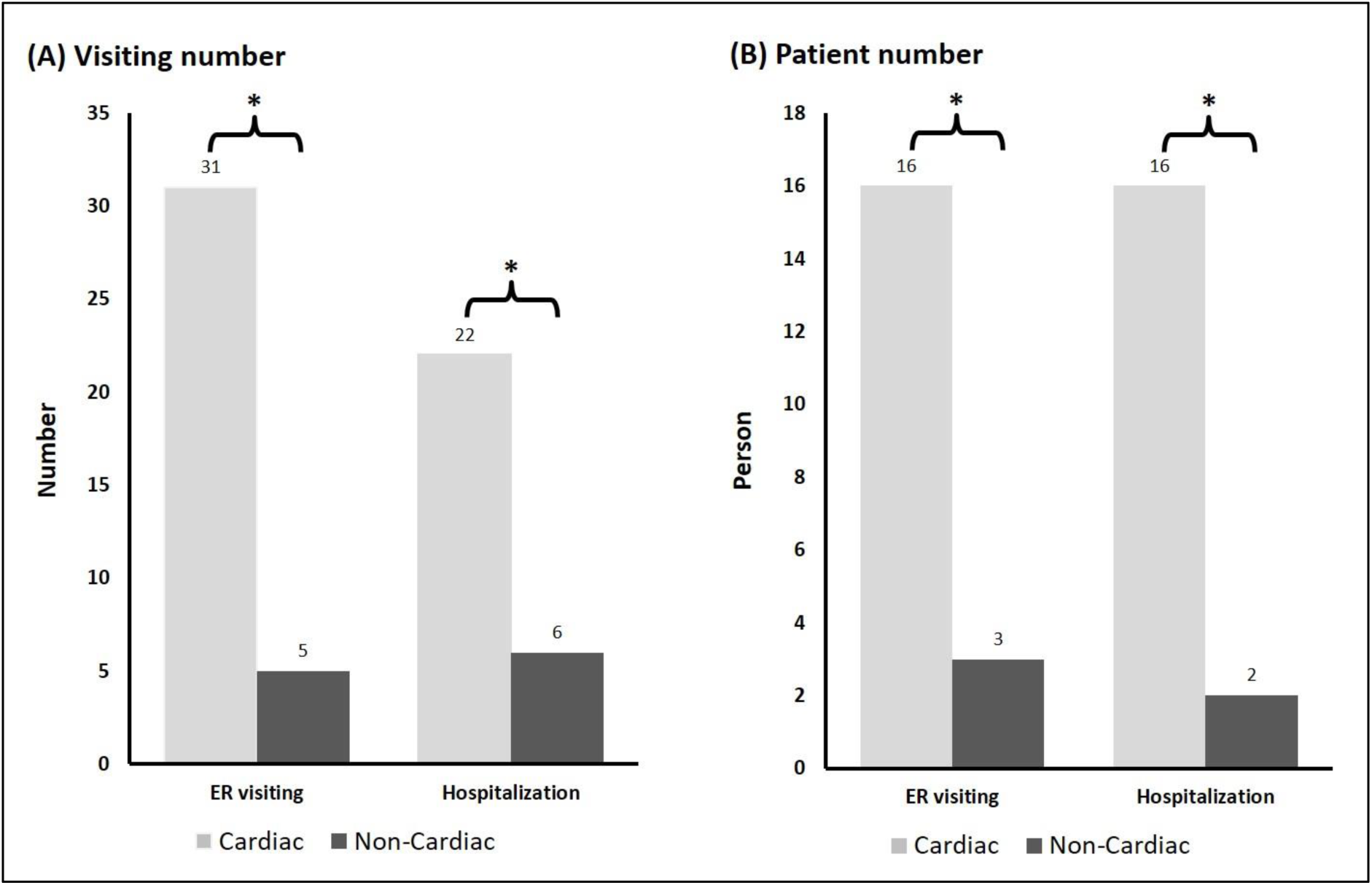
The visiting and patient numbers of ER visiting and hospitalization between the cardiac and non-cardiac groups. (A) Visiting number (B) Patient number * p < 0.05

**Table 2.** The requirement of medical service after hospital discharge.

|  | **Overall**<br>**n = 71** | **Cardiac**<br>**n=58** | **Non-Cardiac**<br>**n=13** | ***p-value*** |
| --- | --- | --- | --- | --- |
| **OPD visiting** | 67 (94.4) | 56 (96.6) | 11 (84.6) | 0.092 |
| First OPD visiting after discharge (day) | 6 (4-9) | 7 (5-9) | 5 (4-7) | 0.101 |
| **ER visiting after hospital discharge** | 23 (32.4) | 20 (34.5) | 3 (23.1) | 0.427 |
| First ER visiting after discharge (day) | 74 (22-286) | 62 (20-147) | 286 (268-298) | 0.100 |
| First ER visiting diagnosis |  |  |  |  |
| Cardiovascular | 8/23 (34.8) | 8 (40.0) | 0 (0) |  |
| Pulmonary | 2/23 (8.7) | 2 (10.0) | 0 (0) |  |
| Infection | 3/23 (13.0) | 2 (10.0) | 1 (33.3) |  |
| Neurologic | 1/23 (4.3) | 1 (5.0) | 0 (0) |  |
| Gastrointestinal | 2/23 (8.7) | 2 (10.0) | 0 (0) |  |
| Endocrine/Metabolism | 2/23 (8.7) | 1 (5.0) | 1 (33.3) |  |
| Miscellaneous | 5/23 (21.7) | 4 (20.0) | 1 (33.3) |  |
| ER diagnosis related to CA cause | 9/23 (39.1) | 8 (40.0) | 1 (33.3) | 0.825 |
| **Hospital admission after hospital discharge** | 22 (31.0) | 18 (31.0) | 4 (30.8) | 0.985 |
| First hospitalization after discharge (day) | 58 (20-257) | 37 (20-166) | 347 (133-905) | 0.173 |
| First hospital admission diagnosis |  |  |  |  |
| Cardiovascular | 13/22 (59.1) | 13 (72.2) | 0 (0) |  |
| Pulmonary | 3/22 (13.6) | 3 (16.7) | 0 (0) |  |
| Infection | 1/22 (4.5) | 0 (0) | 1 (25.0) |  |
| Neurologic | 1/22 (4.5) | 1 (5.6) | 0 (0) |  |
| Endocrine/Metabolism | 2/22 (9.1) | 1 (5.6) | 1 (25.0) |  |
| Autoimmune | 1/22 (4.5) | 0 (0) | 1 (25.0) |  |
| Miscellaneous | 1/22 (4.5) | 0 (0) | 1 (25.0) |  |
| Hospitalization diagnosis related to CA cause | 14/22 (63.6) | 13 (72.2) | 1 (25.0) | 0.076 |
| DAOH from discharge to 1 year (day) | 361 (351-363) | 362 (352-363) | 344 (326-362) | 0.247 |

Data presented as no. (%) or as median (IQR).
ER= emergency room; CA= cardiac arrest; OPD= outpatient department; DAOH= days alive
and out of the hospital

Table 3 presents the work status, neurological function, and quality of life of patients at 3 months and 1 year after hospital discharge. Regarding work status, an increased number of patients were engaged in paid work at 1 year compared with that at 3 months (46.5% vs. 25.4%, p<0.05). No significant difference in neurological outcomes was observed between these two time points. In the EQ-5D-5L assessment, the number of patients with more than moderate impairment significantly decreased from 3 months to 1 year in all domains, except for anxiety/depression. In the WHOQOL-BREF assessment, the scores of all domains were low at 3 months. However, a significant increase in the scores of all domains was observed at 1 year (Supplementary Table 3). Compared with the non-cardiac group, at 3 months, a higher proportion of individuals in the cardiac group were able to return to work (31.0% vs. 0%, p=0.020). Moreover, the individuals in the cardiac group scored higher for general health and psychological well-being than the non-cardiac group at 3 months. Both the EQ-5D-5L and WHOQOL-BREF assessments revealed improvements within the 1-year follow-up period in the cardiac and non-cardiac groups, and the differences between the groups disappeared at 1 year (Table 4).

**Table 3.** Work, neurological recovery and life quality after hospital discharge.

|  | Overall (n = 71) |  | *p-value* |
| --- | --- | --- | --- |
|  | 3 months | 1 year |  |
| **Work** | 18 (25.4) | 33 (46.5) | 0.009 |
| **Favorable neurological outcome** |  |  |  |
| CPC=1 | 63 (88.7) | 66 (93.0) | 0.383 |
| CPC=2 | 6 (8.5) | 3 (4.2) | 0.301 |
| **EQ-5D-5L (more than moderate impaired)** |  |  |  |
| Mobility | 53 (74.6) | 17 (23.9) | <0.001 |
| Self-care | 53 (74.6) | 18 (25.4) | <0.001 |
| Usual activities | 53 (74.6) | 17 (23.9) | <0.001 |
| Pain/discomfort | 52 (73.2) | 18 (25.4) | <0.001 |
| Anxiety/depression | 22 (31.0) | 10 (14.1) | 0.159 |
| Health state | 48 (67.6) | 11 (15.5) | <0.001 |
| **WHOQOL-BREF(score range)** |  |  |  |
| Overall QOL (1–5) | 3 (2-4) | 4 (4-5) | <0.001 |
| General health (1–5) | 3 (2-4) | 4 (4-5) | <0.001 |
| Physical (4-20) | 11.4 (9.1-14.3) | 16.0 (13.7-19.4) | <0.001 |
| Psychological (4-20) | 12.0 (10.7-14.7) | 16.0 (14.0-18.0) | <0.001 |
| Social relationships (4–20) | 13.0 (11.0-16.0) | 16.0 (13.0-17.0) | <0.001 |
| Environment (4-20) | 14.7 (12.0-16.0) | 16.4 (15.6-18.7) | <0.001 |

Data presented as no. (%) or as median (IQR).
CPC= Cerebral Performance Category; EQ-5D-5L= EuroQol 5-Dimension, 5-Level Health
Scale; QOL= Quality of Life; WHOQOL-BREF= World Health Organization Quality of Life-Brief

**Table 4.**
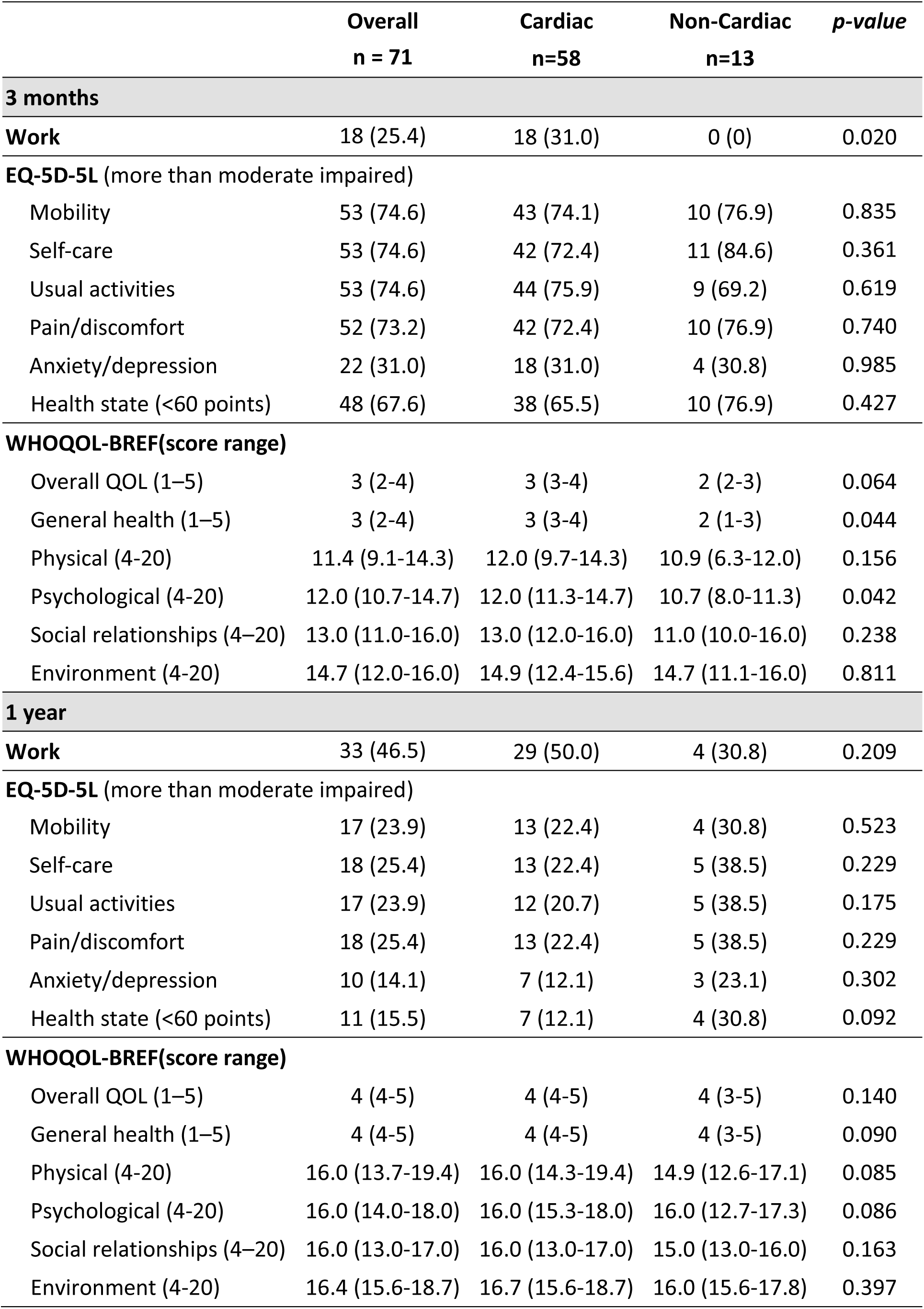
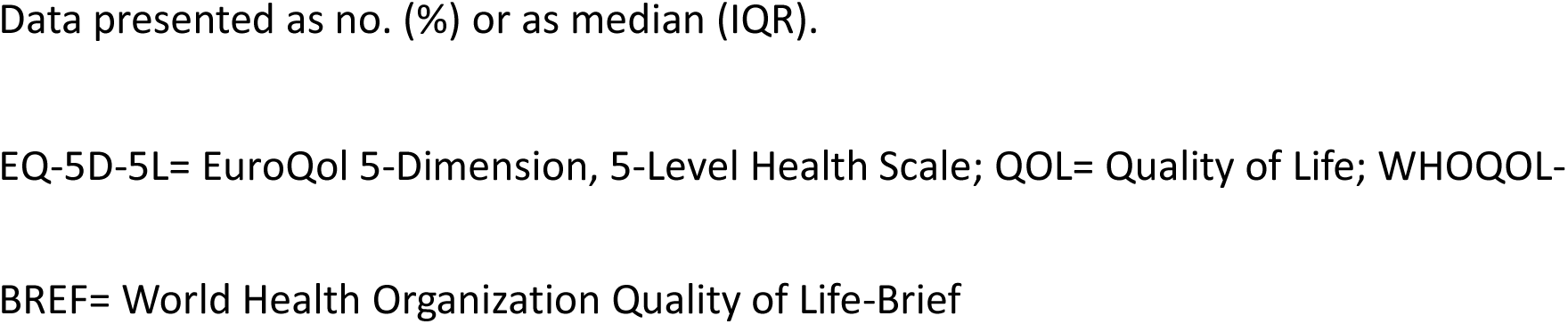
Work and quality of life between the cardiac and non-cardiac groups.

## Discussion

The study retrospectively investigated the medical support requirements, work status, neurological function, and quality of life of cardiac arrest survivors through telephone survey. The results revealed that a considerable number of patients had visited the ER and were readmitted to the hospital within 1 year after discharge. Moreover, at 3 months after discharge, the work status of the cardiac arrest survivors was low, and their quality of life was poor across all aspects measured. However, 1 year after discharge, their work status and quality of life were significantly improved.

In the current study, approximately half of the enrolled cardiac arrest survivors required active medical support within 1 year after discharge, including visits to the ER and hospitalizations. Moreover, one-third of ER visits and half of hospitalizations occurred within 90 days after discharge. Identifying patients who may require active medical support is crucial for providing timely and appropriate care.^9^ In the current study, cardiovascular diagnoses were the leading reasons for medical support after discharge, with most hospitalized patients having diagnoses related to the cause of cardiac arrest. The findings indicate the importance of conducting thorough cardiovascular evaluations and implementing rehabilitation strategies for cardiac arrest survivors after hospital discharge. Mion M. et al.^12^ proposed comprehensive post-discharge care guidelines, emphasizing cardiac rehabilitation, psychological support, lifestyle changes including physical activity and exercise training, and long-term monitoring.

In the current study, the cardiac group exhibited higher work status and quality of life than the non-cardiac group at 3 months, but the difference disappeared at 1 year. This finding indicates that individualized medical and paramedical support after hospital discharge may be necessary.

According to the Victorian Ambulance Cardiac Arrest Registry, 650 of 884 cardiac arrest survivors (73.5%) returned to work during the 12-month follow-up period. This increased rate of return to work was attributable to the favorable HRQoL.^21^ By contrast, in the current study, the number of patients who returned to work within 1 year after hospital discharge was considerably lower than that reported in Western countries.^21^, ^22^ This may be attributed to the lack of comprehensive rehabilitation programs and cultural differences. Similar to the findings obtained using the Victorian Ambulance Cardiac Arrest Registry, the results of the current study demonstrated improvements in work status and quality of life from 3 months to 1 year after discharge. Additional studies are necessary to elucidate the relationship between quality of life and work status in cardiac arrest survivors and to identify the factors that may hinder their return to work.

Schluep M. et al. demonstrated an improvement in the quality of life among cardiac arrest survivors from 3 months to 1 year after hospital discharge, as assessed using the visual analog scale of the EQ-5D-5L.^23^ Posttraumatic symptoms are prevalent in cardiac arrest survivors, and they can persist even years after the initial cardiac arrest event. These symptoms are associated with poorer quality of life in both survivors and informal caregivers.^24^ Notably, in the current study, patients rated psychological demand as considerably lower than physical demand in the EQ-5D-5L assessment, but not in the WHOQoL-BREF assessment. This difference may be attributed to the simplicity and directness of the EQ-5D-5L, which might lead some patients to hesitate or feel embarrassed when expressing their concerns and frustrations. By contrast, the WHOQoL-BREF has multiple and mixed questions, potentially providing a more comfortable and comprehensive platform for patients to express their feelings. This suggests that using a suitable questionnaire is critical for revealing the true condition and experiences of patients. Specific psychometric questionnaire tailored to the needs of cardiac arrest survivors may be required in future investigations to provide a more in-depth understanding of their psychological well-being and quality of life. Early and prompt delivery of interventions can improve the quality of life for cardiac arrest survivors, leading to improvements in their overall emotional states and alleviation of anxiety.^22^ Establishing a comprehensive follow-up protocol and implementing effective health policies for cardiac arrest survivors are crucial.

The current study has several limitations. First, the questionnaire survey was conducted retrospectively; the results may be affected by the memory recall bias among participants. Patients may also tend to underestimate or overestimate their health status. Second, the majority of patients included in the study had a cardiac origin of cardiac arrest, which may have resulted in a more favorable prognosis and recovery compared with patients with non-cardiac causes of arrest. Therefore, the findings may not be generalizable to other populations, particularly those with non-cardiac causes of cardiac arrest. Third, the study could not retrospectively evaluate intelligence and cognitive function. Therefore, the status and changes in the patient’s intelligence and logical thinking over time could not be assessed. Future studies should consider evaluating intelligence and logical thinking. Finally, the study excluded individuals who did not survive the 1-year follow-up period and those who could not be contacted or who declined participation in the study. This exclusion may result in potential selection bias, as these individuals may represent a group in need of even more assistance and support than the patients enrolled in the study.

## Conclusion

This study demonstrated that within 1 year after hospital discharge, a large portion of cardiac arrest survivors require medical support, and that their work status and quality of life continually improved within the first year.

## Disclosures

The authors declare no conflicts of interest for this study.

## Data Availability

No special.

## Notes

### Competing Interest Statement

The authors have declared no competing interest.

### Author Declarations

The study protocol was approved by the Institutional Review Board of NTUH (202107138RIND).

## References

1. Myat A, Song KJ, Rea T. Out-of-hospital cardiac arrest: Current concepts. Lancet. 2018;391:970–979

2. Panchal AR, Bartos JA, Cabanas JG, Donnino MW, Drennan IR, Hirsch KG, Kudenchuk PJ, Kurz MC, Lavonas EJ, Morley PT, et al. Part 3: Adult basic and advanced life support: 2020 american heart association guidelines for cardiopulmonary resuscitation and emergency cardiovascular care. Circulation. 2020;142:S366–S468

3. Nolan JP, Sandroni C, Bottiger BW, Cariou A, Cronberg T, Friberg H, Genbrugge C, Haywood K, Lilja G, Moulaert VRM, et al. European resuscitation council and european society of intensive care medicine guidelines 2021: Post-resuscitation care. Intensive Care Med. 2021;47:369–421

4. Dankiewicz J, Cronberg T, Lilja G, Jakobsen JC, Levin H, Ullen S, Rylander C, Wise MP, Oddo M, Cariou A, et al. Hypothermia versus normothermia after out-of-hospital cardiac arrest. N Engl J Med. 2021;384:2283–2294

5. Merchant RM, Topjian AA, Panchal AR, Cheng A, Aziz K, Berg KM, Lavonas EJ, Magid DJ, Adult B, Advanced Life Support PB, et al. Part 1: Executive summary: 2020 american heart association guidelines for cardiopulmonary resuscitation and emergency cardiovascular care. Circulation. 2020;142:S337–S357

6. Rey JR, Caro-Codon J, Rodriguez Sotelo L, Lopez-de-Sa E, Rosillo SO, Gonzalez Fernandez O, Fernandez de Bobadilla J, Iniesta AM, Pena Conde L, Antorrena Miranda I, et al. Long term clinical outcomes in survivors after out-of-hospital cardiac arrest. Eur J Intern Med. 2020;74:49–54

7. Lilja G, Nilsson G, Nielsen N, Friberg H, Hassager C, Koopmans M, Kuiper M, Martini A, Mellinghoff J, Pelosi P, et al. Anxiety and depression among out-of-hospital cardiac arrest survivors. Resuscitation. 2015;97:68–75

8. Lilja G, Nielsen N, Bro-Jeppesen J, Dunford H, Friberg H, Hofgren C, Horn J, Insorsi A, Kjaergaard J, Nilsson F, et al. Return to work and participation in society after out-of-hospital cardiac arrest. Circ Cardiovasc Qual Outcomes. 2018;11:e003566

9. Djarv T, Bremer A, Herlitz J, Israelsson J, Cronberg T, Lilja G, Rawshani A, Arestedt K. Health-related quality of life after surviving an out-of-hospital compared to an in-hospital cardiac arrest: A swedish population-based registry study. Resuscitation. 2020;151:77–84

10. Lee J, Cho Y, Oh J, Kang H, Lim TH, Ko BS, Yoo KH, Lee SH. Analysis of anxiety or depression and long-term mortality among survivors of out-of-hospital cardiac arrest. JAMA Netw Open. 2023;6:e237809

11. Sawyer KN, Camp-Rogers TR, Kotini-Shah P, Del Rios M, Gossip MR, Moitra VK, Haywood KL, Dougherty CM, Lubitz SA, Rabinstein AA, et al. Sudden cardiac arrest survivorship: A scientific statement from the american heart association. Circulation. 2020;141:e654–e685

12. Mion M, Simpson R, Johnson T, Oriolo V, Gudde E, Rees P, Quinn T, Vopelius-Feldt VJ, Gallagher S, Mozid A, et al. British cardiovascular intervention society consensus position statement on out-of-hospital cardiac arrest 2: Post-discharge rehabilitation. Interv Cardiol. 2022;17:e19

13. Devlin N, Pickard S, Busschbach J. The development of the eq-5d-5l and its value sets. In: Devlin N, Roudijk B, Ludwig K, eds. Value sets for eq-5d-5l: A compendium, comparative review & user guide. Cham (CH); 2022:1–12.

14. Skevington SM, Epton T. How will the sustainable development goals deliver changes in well-being? A systematic review and meta-analysis to investigate whether whoqol-bref scores respond to change. BMJ Glob Health. 2018;3:e000609

15. Ling DA, Huang CH, Chen WJ, Chuang PY, Chang WT, Sung CW, Chen WT, Ong HN, Tsai MS. Impact of protocolized postarrest care with targeted temperature management on the outcomes of cardiac arrest survivors without temperature management. Ann Med. 2022;54:63–70

16. Chen WT, Tsai MS, Huang CH, Chang WT, Chen WJ. Protocolized post-cardiac arrest care with targeted temperature management. Acta Cardiol Sin. 2022;38:391–399

17. Sanchis J, Bueno H, Minana G, Guerrero C, Marti D, Martinez-Selles M, Dominguez-Perez L, Diez-Villanueva P, Barrabes JA, Marin F, et al. Effect of routine invasive vs conservative strategy in older adults with frailty and non-st-segment elevation acute myocardial infarction: A randomized clinical trial. JAMA Intern Med. 2023;183:407–415

18. Devlin NJ, Brooks R. Eq-5d and the euroqol group: Past, present and future. Appl Health Econ Health Policy. 2017;15:127–137

19. Yao G, Chung CW, Yu CF, Wang JD. Development and verification of validity and reliability of the whoqol-bref taiwan version. J Formos Med Assoc. 2002;101:342–351

20. Health organization. (1998). Programme on mental health : Whoqol user manual, 2012 revision. World health organization. (accessed may 10, 2023, at: https://apps.Who.Int/iris/bitstream/handle/10665/77932/who_his_hsi_rev.2012.03_eng.Pdf?Sequence=1&isallowed=y).

21. Kearney J, Dyson K, Andrew E, Bernard S, Smith K. Factors associated with return to work among survivors of out-of-hospital cardiac arrest. Resuscitation. 2020;146:203–212

22. Moulaert VR, van Heugten CM, Winkens B, Bakx WG, de Krom MC, Gorgels TP, Wade DT, Verbunt JA. Early neurologically-focused follow-up after cardiac arrest improves quality of life at one year: A randomised controlled trial. Int J Cardiol. 2015;193:8–16

23. Schluep M, Endeman H, Gravesteijn BY, Kuijs C, Blans MJ, van den Bogaard B, Van Gemert A, Hukshorn CJ, van der Meer BJM, Knook AHM, et al. In-depth assessment of health-related quality of life after in-hospital cardiac arrest. J Crit Care. 2022;68:22–30

24. Presciutti A, Newman MM, Grigsby J, Vranceanu AM, Shaffer JA, Perman SM. Associations between posttraumatic stress symptoms and quality of life in cardiac arrest survivors and informal caregivers: A pilot survey study. Resusc Plus. 2021;5:100085

